# I’m alone but not lonely. U-shaped pattern of perceived loneliness during the COVID-19 pandemic in the UK and Greece

**DOI:** 10.1101/2020.11.26.20239103

**Authors:** Alessandro Carollo, Andrea Bizzego, Giulio Gabrieli, Keri Ka-Yee Wong, Adrian Raine, Gianluca Esposito

## Abstract

Many countries have adopted lengthy lockdown measures to mitigate the spreading of the COVID-19 virus. In this study, we train a RandomForest model using 10 variables quantifying individuals’ living environment, physical and mental health statuses to predict how long each of the UK participants (N=382) had been in lockdown. Self-perceived loneliness was found to be the most important variable predicting time in lockdown and, therefore, the aspect most influenced by the time the participant spent in lockdown. Subsequent statistical analysis showed a significant U-shaped curve for the levels of perceived loneliness (p<0.012), specifically decreasing during the 4th and 5th lockdown weeks. The same pattern was found on data from Greek citizens (N=129, p<0.041). These results suggest that lockdown measures may have affected how people evaluated their social support while in lockdown, leading to a decreased sense of loneliness. Implications of this study should be reflected on policies and countermeasures to current and future pandemics.

**State of relevance:** This study aims to inform policies for the current and/or future pandemics, particularly those involving lockdown restrictions. It highlights that self-perceived loneliness was the trait most affected by the time spent in lockdown: data show that the very first period of lockdown was characterised by a decrease in levels of perceived loneliness. The machine learning approach adopted and the statistical validation on two different Western European countries ensure that the uncovered pattern is substantial. This result highlights the dissociation between objective social support and perceived loneliness: initially, restrictions may have triggered better social behaviours among communities or increased the level of gratitude for the social support people have always received. The short duration of these desirable effects suggests that measures and campaigns promoting better social support strategies could be potentially effective, even in social isolation, to keep the levels of perceived loneliness low.

## Introduction

The 2019 SARS-CoV-2 (COVID-19) outbreak was declared as a pandemic by the World Health Organisation (WHO) on 11 March 2020. The number of positive cases worldwide at the time was 179,111 and deaths, 7,426 (Organization, 2020b). Globally, the months that followed saw a surge in the number of deaths and infection rates, which put further strain on the sanitary and economical balance of several countries. Fast forward to September 13th 2020, the total number of confirmed COVID-19 cases at 28,637,952 and 917,417 deaths have since been recorded (Organization, 2020a).

Expert concern for the mental health consequences of the current pandemic stems from the evidence that was obtained during smaller epidemics, such as SARS (severe acute respiratory syndrome), MERS (Middle East respiratory syndrome-related coronavirus), H1N1, and Ebola. From these previous health emergencies, short- and long-term effects on the healthcare workers’ mental health, such as post-traumatic stress disorder (PTSD) (S. M. Lee et al., 2018; Maunder et al., 2006), depression (A. M. Lee et al., 2007; Liu et al., 2012), anxiety (A. M. Lee et al., 2007), stress and burnout (Maunder et al., 2006) symptoms were common (Preti et al., 2020). There is evidence that healthcare workers are distressed from the epidemics, during and after emergencies, and that these effects also extend to the general population in the form of severe anxiety, post-traumatic stress disorder, depression and increased rates of substance abuse (Brooks et al., 2020; Haider et al., 2020; Lau et al., 2005; Main et al., 2011; Mak et al., 2009; Zhang & Ma, 2020). Although some promising results from vaccine trials are starting to emerge, the novel and highly infective virus continues to force governments around the world to limit people’s movements and, in some cases, re-adapt lockdown restrictions once again, as in the case of the UK on September 22nd, 2020. Closing schools and universities, shutting non-essential businesses, enforcing working from home policies and online teaching, struggling with financial difficulties and leaving the house only for necessities have fuelled genuine and perceived health threats that have rapidly become ubiquitous for large populations worldwide. While restrictions have helped flatten the infection curve, legitimate concerns about the physical and mental health consequences have been raised. As such, this pandemic, as an extreme global stressor, has provided an unprecedented opportunity for researchers to investigate how several aspects of our personal life, and specifically our mental health, are affected by prolonged isolation and restrictions. Social isolation is one known threat to mental and physical well-being (Hall-Lande et al., 2007; Valtorta et al., 2016) and an established risk factor for mortality (Alcaraz et al., 2019; Cacioppo et al., 2015; Holt-Lunstad et al., 2015). Social isolation is associated with poor sleep quality (Friedman, 2011) and with an increased risk of cognitive decline (Barnes et al., 2004). The fact that our perception of self is ingrained in the social comparison with others (Festinger, 1954) ssuggests that social isolation may not be an ideal situation for the development of our identity either. Latest COVID-19 studies of the first weeks of lockdown have already documented psychological distress, such as depression, anxiety, post-traumatic stress and insomnia in Italy (Castelli et al., 2020; Gualano et al., 2020; Rossi et al., 2020) and China (Ahmed et al., 2020; Wang et al., 2020), the two countries most severely hit by COVID-19 at the beginning of the pandemic, as well as Austria (Pieh et al., 2020) and Switzerland (Elmer et al., 2020). In fact, with some preliminary results on COVID-19 restrictions, this paper aims to add a piece of knowledge to the existing literature to provide a scientific contribution and help governments in the design of future possible lockdowns. Against this backdrop of existing psychological consequences from lockdown, this study focuses on the physical and psychological constructs that best predict the time spent in lockdown (TIL).

## Methods

### Questionnaire

A 20-minute online survey (available in 7 languages) was administered through the website www.GlobalCOVIDStudy.com between 17 April 2020 and 10 July 2020 to participants aged 18 years and above who had access to the survey link. This was distributed using various social media channels (email, LinkedIn, Whatsapp, Instagram, Facebook and Reddit). The survey was designed by the Global COVID Study team in order to explore participants’ moods and behaviours. The battery of questionnaires consisted of 359 questions assessing 13 main domains: social suspicions, schizotypal traits, physical health, sleep quality, aggression, empathy, anxiety, depression, worries and stress, loneliness, parenting style, Special Educational Needs and demographic information (see https://osf.io/fe8q7 for more details). The study was approved by the UCL Ethics Committee (REC 1331).

### Participants

Participants for the study were recruited through convenience sampling and, eventually, a total of 2,276 people (aged 18 and above) from 66 countries completed the survey during lockdown. Respondents who did not give consent to treat their data (N = 32), with incomplete (N = 712) or missing data (N = 294), or who could not complete the survey within two days from their enrollment (N = 76) were excluded. To train the Random Forest, we chose not to consider the participants who took more than one day because the process required the model to find patterns of dependency between the features and the amount of time spent in lockdown. Considering the fact that, in our hypothesis, the time in lockdown played a role in determining the variability of the selected features, by considering only the participants who completed the survey within the same day, we aimed at reducing possible confounds. Furthermore, participants who completed the survey from a country that was different from the one they were a resident of were excluded from the study (N = 132). Considering the variety of lockdown measures across the world, this criterion was adopted in order to reduce possible confounds given by the type of restrictions adopted by individual countries. Another possible confound came from the fact that not all the governments decided to adopt lockdown restrictions against the pandemic and, when they did, different countries entered the lockdown on different dates. For these reasons, among the countries that adopted these restrictions, the new variable “Weeks in lockdown” - the time of survey completion - was computed for each participant. Thus, participants were grouped and compared regardless of the specific date in which their countries decided to adopt restrictions, but uniquely by the amount of time spent in lockdown. Within this pool of data, the UK and Greece samples were selected for the analysis conducted in this study. Three main reasons drove the choice of using data from these two countries: a) the sample sizes (>100 cases); b) the existence of a clear date of lockdown beginning; and c) the same time span (weeks in lockdown) covered. To maintain the coverage on the same time period, UK participants that completed the survey after week 9 of lockdown were excluded from the study (N = 40). To summarise, the UK sample consisted of 382 participants (Gender: Female = 302, Male = 71, Non-binary = 4, Prefer not to say = 2, Self-identified = 3; Age: mean = 37.18; SD = 13.15), while the Greek sample counted 129 participants (Gender: Female = 92, Male = 37; Age: Mean = 36.08, SD = 10.79) (see Table 1).

**Table 1.**
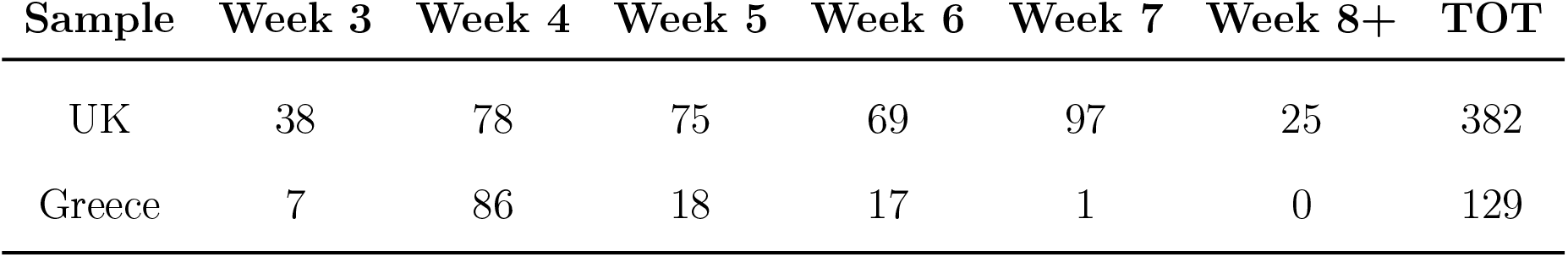
Distribution of participants from the UK and Greece by week.

### Data Analysis

From the dataset, 10 variables capturing participants’ living environment, mental and physical health were selected (see Table 2). This procedure aimed to remove the noise in the dataset by discarding the non-informative variables. All the variables for which scores could not show a variation caused by the amount of time in lockdown were excluded. Some examples of these variables are gender, ethnicity, dimensions of one’s house, and other similar variables. Moreover, considering the criteria for which data from participants could potentially have at least one NA data were not considered for the analysis, the next step of the selection was to optimise the number of participants by not considering the variables that had a large amount of NA data.

**Table 2.**
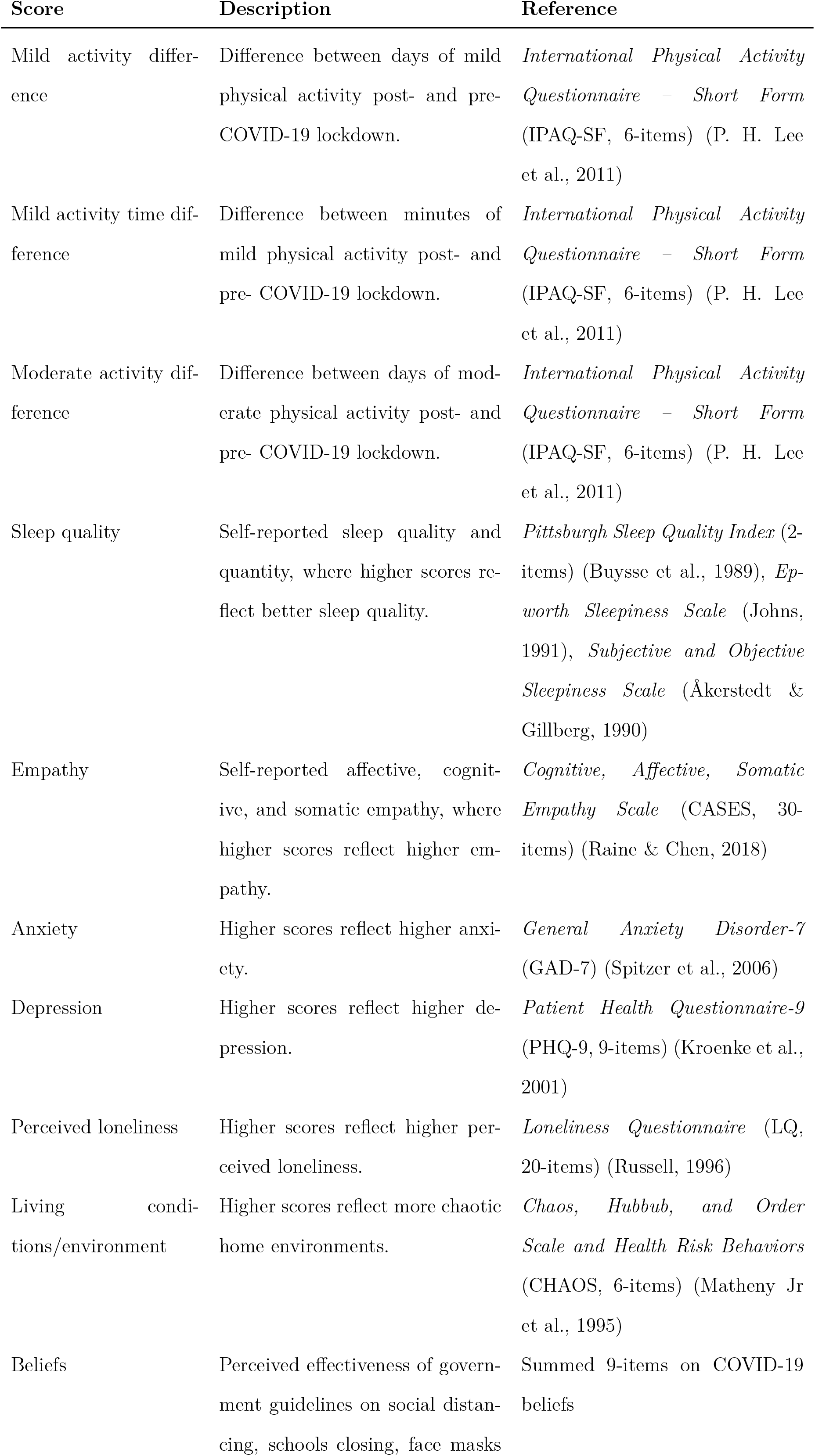
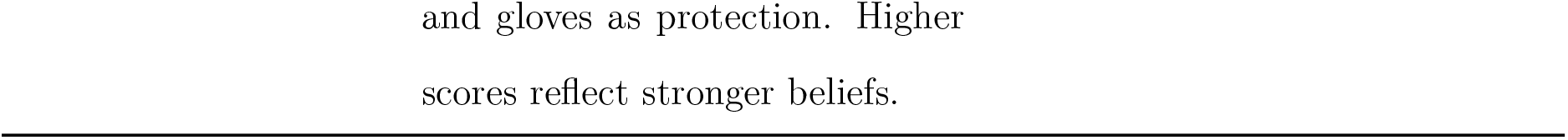
Scores that are computed to quantify participants’ mental and physical health and living environment during lockdown.

In order to investigate the role of time spent in lockdown on modulating the effects of lockdown restrictions (study pre-registration: https://osf.io/8xtbq), the study consisted of two parts. In the first, we aimed at understanding which aspect was most sensitive to time. In other words, which part of our physical and mental life was affected the most by the time in lockdown. In the second, the interest was on how this most sensitive aspect was modulated by the time.

#### Identification of the most influential variable

Without any available literature to guide our hypothesis in identifying the variable that is most influenced by TIL, we adopted a data-driven Machine Learning approach where a RandomForest (Breiman, 2001) regression model was trained to predict the week in which each participant completed the survey, based on the total scores of the 10 selected variables. The model creates an ensemble of decision trees based on the predictive information of the input variables. The performance of the model was evaluated based on Mean Squared Error (MSE). The data used to train the RandomForest model were those of the 382 UK residents who were in the UK at the time of participation in this study. Initially, the dataset was partitioned into train (75% of participants) and test (25% of participants). The training process was repeated and evaluated several times on different randomized folds of the train dataset to optimize the number of decision trees and rank the variables based on their importance. A Borda count (Jurman et al., 2012) was then computed on the rankings of variables obtained from each training iteration to identify the most important variable to predict the week of survey completion. The optimal number of decision trees that emerged from the training was 50. The final model, with the optimal number of trees, was then trained on the whole train partition and evaluated on the test partition. The adopted training scheme is standardized and was derived from bioinformatics applications that are used to identify clinical biomarkers from genetic data (Consortium et al., 2010).

#### Statistical validation

In the second part of the study, we used a Kruskal-Wallis test to assess whether the most important variable (identified by the RandomForest model) significantly changes during the lockdown from weeks 3 to 7. In case of significant results, we adopted post-hoc Kruskal-Wallis tests to compare pairwise the 3rd week with the 4th to 7th weeks. The Bonferroni method was used to correct the significance level for multiple comparisons. In conducting statistical analyses, we first focused on the same set of participants used to train the RandomForest model, then we validated results on the dataset of participants from Greece.

## Results

MSE on the training and the test partitions was 1.33 and 1.94 respectively and the feature with the highest importance was perceived loneliness (see Figure 1) Notably, scores of perceived loneliness decreased during weeks 3 to 5 after lockdown and subsequently increased in the following weeks, returning to the initial values (see Figure 2). The Kruskal-Wallis test on the data of UK participants from the 3rd to 7th week confirmed that at least one week was statistically different from the others (H=12.86, p=0.012). We then compared the 4th, 5th, 6th and 7th week with the 3rd. Significant differences were found for the 4th (H=11.360, p=0.001) and 5th (H=7.077, p=0.008) week, but not for the 6th (H=4.011, p=0.045) and the 7th (H=0.368, p=0.544) week. The same procedure was repeated on participants from Greece, focusing only on the 3rd to 6th weeks, as only one participant completed the survey during the 7th week. The results confirmed that perceived loneliness significantly changes over weeks 3 to 6 (H=8.27, p=0.041), with a significant difference between the 3rd and the 5th weeks (H=7.6, p=0.006). The difference between the 3rd and the 4th weeks (H = 3.87, p=0.049) failed to survive the Bonferroni correction. No difference was found between the 3rd and the 6th week (H=0.68, p=0.408).

**Figure 1.**
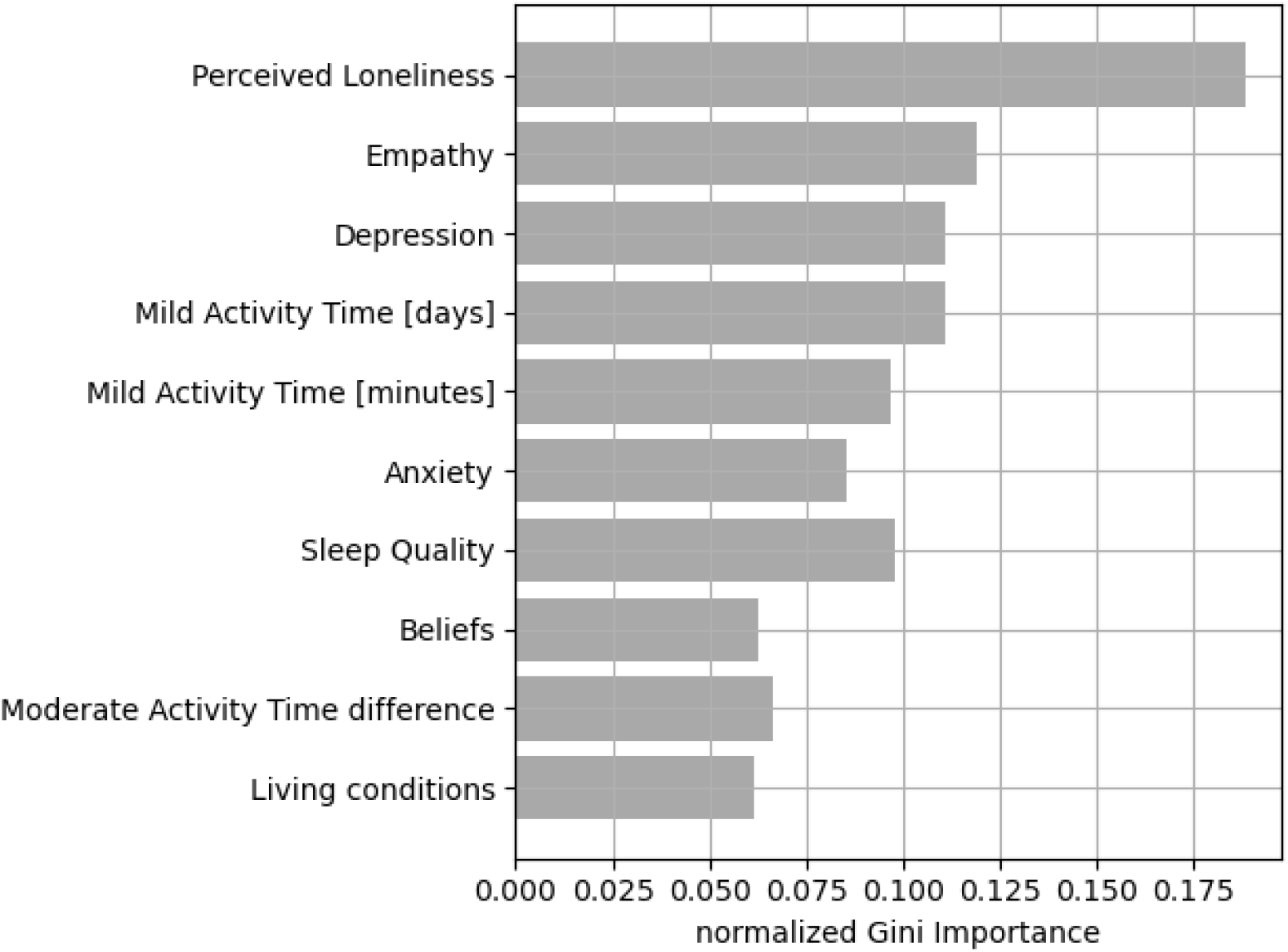
Average importance of the selected variables.

**Figure 2.**
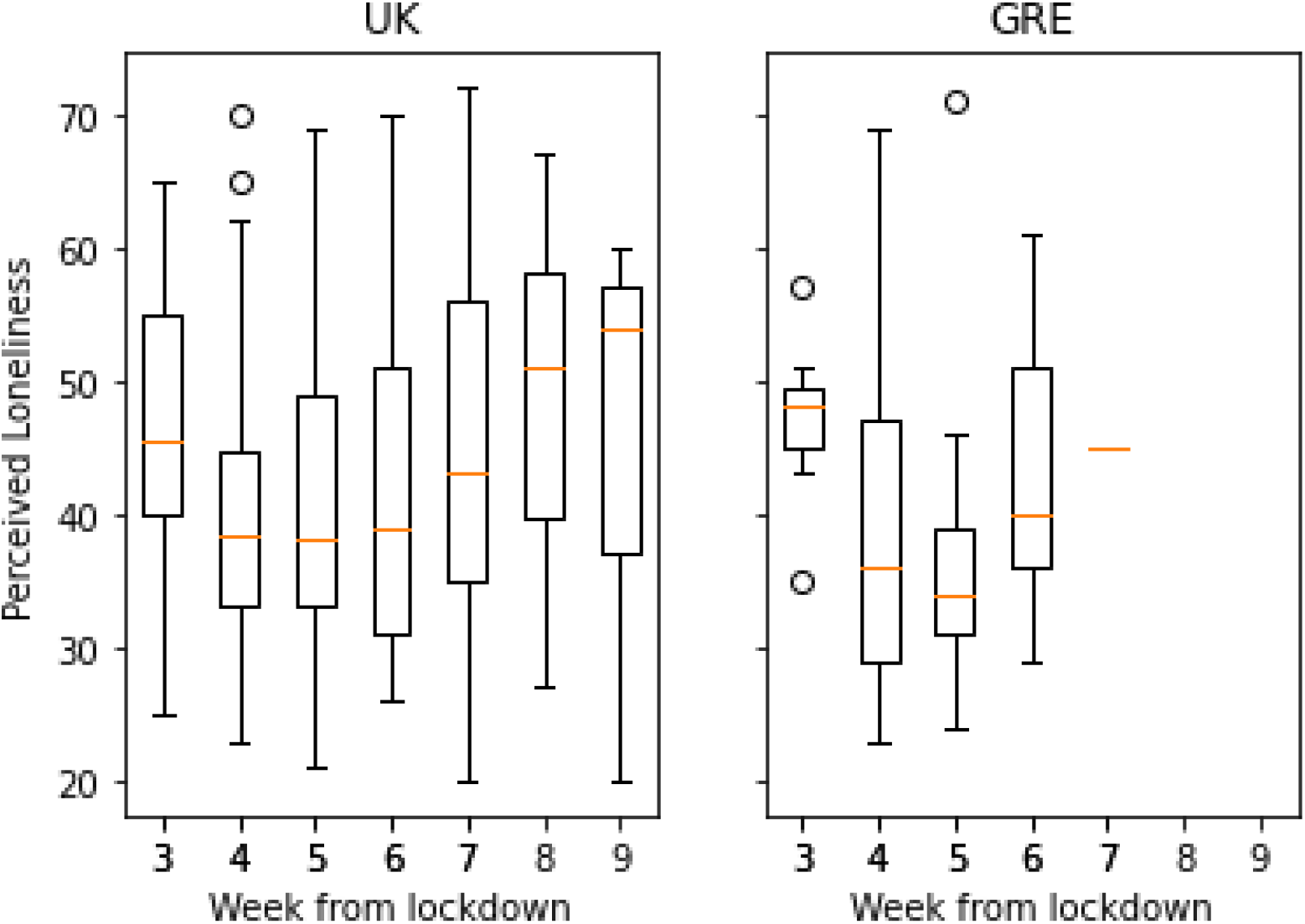
Distribution of perceived loneliness scores for each week for participants from the UK (left) and Greece (right).

## Discussion

The aim of this study was to assess the impact of lockdown restrictions on people’s mental and physical health. Although we adopted a rigorous methodology to train the predictive model, the achieved performances on train and test partitions are low: this outcome reflects the complexity of the psycho-social mechanisms that were at play during the lockdown period. While the questionnaires aimed at quantifying a broad range of the aspects of interest, other aspects may not have been observed. Additionally, this study investigated the temporal variations of these mechanisms, whose effects on the observed variables might be even more difficult to identify. That said, one advantage of the machine learning approach is that it permits the identification of variables that are more sensitive to the time spent in lockdown, rather than a focus on predictive capability.

The low performance does not affect the reliability of the ranking of the variables, which identified perceived loneliness as the most sensitive variable. Perceived loneliness in the UK decreased during the first 4-5 weeks after the start of the lockdown, before returning to initial values afterwards. The pattern was replicated in the Greek sample, albeit in a smaller group of participants. This confirms that perceived loneliness does capture a sensitive decrease during weeks 4-5 since the start of lockdown.

These results are somewhat surprising. In the emerging literature about COVID-19, the number of friends and one’s social support seem to play a protective role against the effect of lockdown on loneliness (Bu et al., 2020a, 2020b). Counterintuitively, from our study it emerged that, even though a large part of the global population was not able to see their close friends, partner and family, levels of perceived loneliness interestingly decreased during the initial period of lockdown. The dissociation between the objective and the subjective degrees of social support has been largely discussed in the existing literature (Cacioppo & Hawkley, 2009; Hawkley et al., 2008). It is believed that perceived social isolation represents a quantitative or (more often) qualitative mismatch between an individual’s need for social support and the subjective evaluation of the social support that is obtained (Hawkley & Capitanio, 2015). In other words, the feeling of loneliness, resulting from the perception of social isolation, seems to depend, more than on an objective condition of isolation, on a cognitive evaluation and perception of the social environment. In the existing literature, the feeling of loneliness emerged to be connected to the concept of Self (Goswick & Jones, 1981), the person’s cognitive functioning (Cacioppo & Hawkley, 2009) and, in general, the mental and physical well-being of the individual (Cacioppo et al., 2015; Holt-Lunstad et al., 2015; Richardson et al., 2017; Valtorta et al., 2016). For instance, lonely people are more likely to suffer from depression (Jackson & Cochran, 1991; Mushtaq et al., 2014), Alzheimer’s disease (Holwerda et al., 2014; Mushtaq et al., 2014), alcoholism (Åkerlind & Hörnquist, 1992; Hawkley & Cacioppo, 2010; Mushtaq et al., 2014), suicide (Mushtaq et al., 2014; Stravynski & Boyer, 2001), personality disorders (Mushtaq et al., 2014) and sleep problems (Mushtaq et al., 2014). It is not yet clear the reason behind the results that emerged from this study, but some hypotheses can be advanced. For instance, considering the definition of loneliness as a mismatch between desired and obtained social support, the decrease of its levels in the first weeks of lockdown could signify that people in that period of time were receiving the desired social support or even more of it in terms of quantity, or that it is higher in quality. On the other hand, loneliness could decrease as a result of a drop in the standards used for evaluating the received social support. In times of danger, this could be an adaptive feature, for it could facilitate behaviours of affiliation among people of the same group. As a matter of fact, facing an external threat has the short-term effect of increasing cohesiveness among the members of a group (Sherif et al., 1961; Staw et al., 1981). The threat of external dangers (such as invaders) has often been used from past and present political leaders in order to rule their countries and to increase the sense of community among the population. It is possible that not only personified external threats like foreigners trying to invade one’s country, but also environmental dangers, such as the COVID-19 pandemic, can directly modulate the degree of cohesiveness among the members of a group, in this case an entire population of a country. As a matter of fact, this increased cohesion could have had a role in the initial decrease in levels of perceived loneliness that we observed among people from the UK and Greece. Even though no certain explanation can be given for understanding the observed patterns of perceived loneliness, the findings of this study support the idea that social isolation (as the objectively low social support) and loneliness (as the subjectively low social support) are different concepts, not necessarily linked to each other, as philosophers in the past centuries have largely pointed out. Having observed that lockdown restrictions have short-term effects on people’s feeling of loneliness and not knowing the real meaning behind these observed patterns, in our opinion, the design of possible future lockdown measures should be accompanied by the consideration of the role played by real and perceived social support for people’s physical and mental well-being.

## Data Availability

Data will be available online after an embargo period

https://rdr.ucl.ac.uk/articles/dataset/I_m_alone_but_not_lonely_U-shaped_pattern_of_perceived_loneliness_during_the_COVID-19_pandemic_in_the_UK_and_Greece/13251893

## Author contribution

Conceptualization: A.B., G.G., K.K.W., G.E.; Data curation: A.C., A.B., G.G., K.K.W.; Data analysis, Data interpretation, Writing: A.C., A.B.; Revision: A.C., A.B., G.G., K.K.W., A.R., G.E.; Supervision: G.E. All authors read and agreed to the published version of the manuscript.

## Conflicts of interest

The authors declare no conflict of interest.

## Ethics

Ethical approval for the COVID-19 Social Study was granted by the UCL Ethics Committee (REC 1331). The study is GDPR compliant.

## Funding

This research is supported by Nanyang Technological University (Singapore) under the NAP-SUG grant (G.E.).

## Notes

### Competing Interest Statement

The authors have declared no competing interest.

